# Defining the Subtypes of Long COVID and Risk Factors for Prolonged Disease

**DOI:** 10.1101/2023.05.19.23290234

**Authors:** Skyler Resendez, Steven H. Brown, H. Sebastian Ruiz, Prahalad Rangan, Jonathan R. Nebeker, Diane Montella, Peter L. Elkin

**Affiliations:** Department of Biomedical Informatics, Jacobs School of Medicine and Biomedical Sciences, University at Buffalo; Department of Veterans Affairs, VA Western New York Healthcare System and VA Research Service; Office of Health Informatics, Department of Veterans Affairs; Department of Biomedical Informatics Vanderbilt University Medical Center; Department of Medicine, University of Utah; Department of Internal Medicine, Jacobs School of Medicine and Biomedical Sciences, University at Buffalo; Faculty of Engineering, University of Southern Denmark

## Abstract

**Importance:** There have been over 759 million confirmed cases of COVID-19 worldwide. A significant portion of these infections will lead to long COVID and its attendant morbidities and costs.

**Objective:** To empirically derive a long COVID case definition consisting of significantly increased signs, symptoms, and diagnoses to support clinical, public health, research, and policy initiatives related to the pandemic.

**Design:** Case-Crossover Population-based study.

**Setting:** Veterans Affairs (VA) medical centers across the United States between January 1, 2020 and August 18, 2022.

**Participants:** 367,148 individuals with positive COVID-19 tests and preexisting ICD-10-CM codes recorded in the VA electronic health record were enrolled.

**Trigger:** SARS-CoV-2 infection documented by positive laboratory test.

**Case Window:** One to seven months following positive COVID testing.

**Main Outcomes and Measures:** We defined signs, symptoms, and diagnoses as being associated with long COVID if they had a novel case frequency of >= 1:1000 and they were significantly increased in our entire cohort after a positive COVID test when compared to case frequencies before COVID testing. We present odds ratios with confidence intervals for long COVID signs, symptoms, and diagnoses, organized by ICD-10-CM functional groups and medical specialty. We used our definition to assess long COVID risk based upon a patient’s demographics, Elixhauser score, vaccination status, and COVID disease severity.

**Results:** We developed a long COVID definition consisting of 323 ICD-10-CM diagnosis codes grouped into 143 ICD-10-CM functional groups that were significantly increased in our 367,148 patient post-COVID population. We define seventeen medical-specialty long COVID subtypes such as cardiology long COVID. COVID-19 positive patients developed signs, symptoms, or diagnoses included in our long COVID definition at a proportion of at least 59.7% (based on all COVID positive patients). Patients with more severe cases of COVID-19 and multiple comorbidities were more likely to develop long COVID.

**Conclusions and Relevance:** An actionable, empirical definition for long COVID can help clinicians screen for and diagnose long COVID, allowing identified patients to be admitted into appropriate monitoring and treatment programs. An actionable long COVID definition can also support public health, research and policy initiatives. COVID patients with low oxygen saturation levels or multiple co-morbidities should be preferentially watched for the development of long COVID.

## Long COVID – Current Knowledge Base and Need

Numerous symptoms are cited as long-term sequela of COVID-19. “The symptoms may affect a number of organ systems, occur in diverse patterns, and frequently get worse after physical or mental activity.”^1^ A 2020 study found that the most common long-term symptoms were fatigue, dyspnea, joint pain, and chest pain.^2^ Others reported gastrointestinal tract disorders correlated with gut microbiome shifts after COVID-19 infection.^3,4^ Cognitive dysfunction, often referred to as brain fog, is a commonly reported long-term symptom. ^5^ Cognitive dysfunction is particularly concerning given evidence that COVID-19 can alter brain structure.^6^ The most common self-reported symptoms documented via a smartphone app were fatigue, headache, dyspnea, and anosmia.^7,8^

Concerningly high long COVID frequencies have been reported. A cohort study from the Netherlands found that approximately one in eight COVID-19 patients developed long-term somatic symptoms.^9^ Another study showed that approximately thirty percent of their cohort reported persistent symptoms, with many experiencing worse health-related quality of life (HRQoL) compared with baseline and negative impacts on at least one activity of daily living.^10^

Long COVID’s impacts extend beyond individual morbidity to include healthcare system and economic consequences. Cutler et. al. noted long COVID resulting in reduced workforce participation (e.g., 44% out of the work force), direct earning losses and worker shortages in service jobs. ^11^ A recent analysis of New York State disability claims trends described in the New York Times found that “71 percent of claimants with long COVID needed continuing medical treatment or were unable to work for six months or more” and opined that long COVID has exacerbated the current US labor shortage.^12,13^

The widespread occurrence of lingering ailments and their impacts on individuals and society make clear the need for a long COVID definition. U.S. public health officials note that we must balance our need for an accurate long COVID definition that includes all afflicted individuals with against our need for interim long COVID definitions to expedite immediate action and mobilization.^14^ In particular, a working definition of long COVID based on routinely collected coded data could support the identification of at risk or undiagnosed patients for monitoring, referral, or therapeutic interventions. In the current study we empirically derive an actionable broad-based long COVID definition to support current clinical, public health, research and policy initiatives related to the pandemic.

## Methods

### Overview

We enrolled Veterans who had laboratory confirmed positive COVID 19 tests. We examined Veterans’ electronic health records for novel International Classification of Diseases, 10th Revision, Clinical Modification (ICD-10-CM) codes between one and seven months after a positive COVID-19 test. We grouped codes with a novel frequency of 1/1000 or greater by diagnosis type creating ICD-10-CM functional groups and performed Chi-square testing with Bonferroni correction to compare diagnosis frequencies before and after a positive COVID test. We defined ICD-10-CM functional groups that significantly increased in frequency as “upregulated” (see Figure 1). We then manually aggregated upregulated ICD-10-CM functional groups into medical specialties to organize our empiric definition of long COVID.

**Figure 1.**
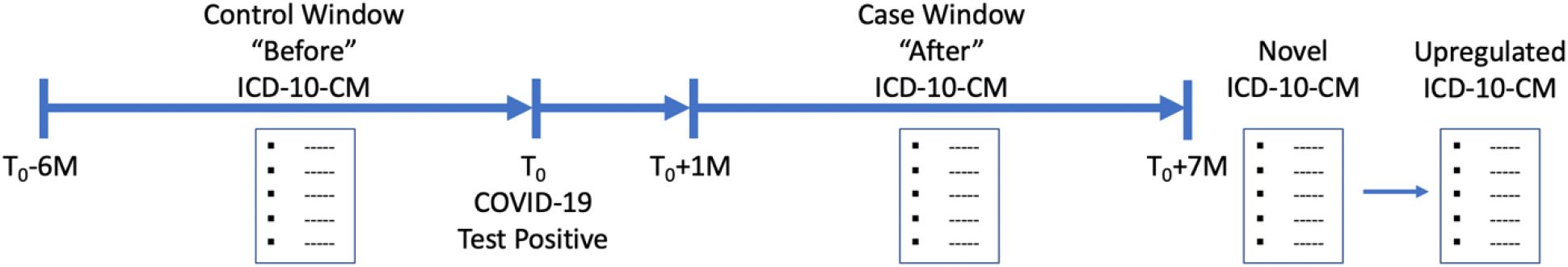
Workflow and data curation in the sequence utilized to generate our long COVID definition. T_0_: Date of positive COVID-19 test. M: Months.

### Population Definition and Data Extraction

We enrolled patients with laboratory confirmed positive COVID-19 studies and followed them for thirteen months (Six months before the COVID 19 test result and seven months after COVID) to create a long COVID definition. We utilized the electronic health records (EHR) of all patients that tested positive for COVID-19 at Veterans Affairs (VA) medical facilities nationally between January 1, 2020 and August 18, 2022. 2,377,720 patients were tested for COVID-19 during this time period.

We applied structured query language (SQL) queries to VA Informatics and Computing Infrastructure (VINCI), Corporate Data Warehouse (CDW) data tables^15^ to generate two diagnosis files for analysis. The first file (“before”) contains a row of retrospectively collected information for each patient and each ICD-10-CM diagnosis assigned to them in the six-month control window prior to their COVID-19 test. The row includes the ICD-10-CM code and its description, a unique patient identifier, the COVID-19 test date, and the calculated number of months between ICD-10-CM code entry and COVID-19 testing. This “before” file contained 14,980,288 observations across 426,970 patients.

We followed the patients for seven months. The second file (“after”) was created seven months after the last patient was enrolled. The after file contained ICD-10-CM codes assigned during the seven months following COVID-19 testing and similar related information as the “before” file. This “after” file contained 15,493,587 observations across 389,677 patients.

We limited analysis to the 367,148 patients that appeared in both the “before” and “after” files to ensure that we had a diagnostic history for each patient and eliminated acute findings by removing all ICD-10-CM codes documented less than a month after the positive COVID test (Figure 1). We used the date of the first positive COVID test for patients with multiple positive tests. Multiple repeating ICD-10-CM codes for a single patient were counted once. We wrote R and python programs to remove all data concerning patients who tested negative for COVID-19, ICD-10-CM codes that were documented less than a month after the positive COVID-19 test, and patients who were not present in both the “before” and “after” files. The methodology used to generate the patient cohort is depicted in Figure 2.

**Figure 2.**
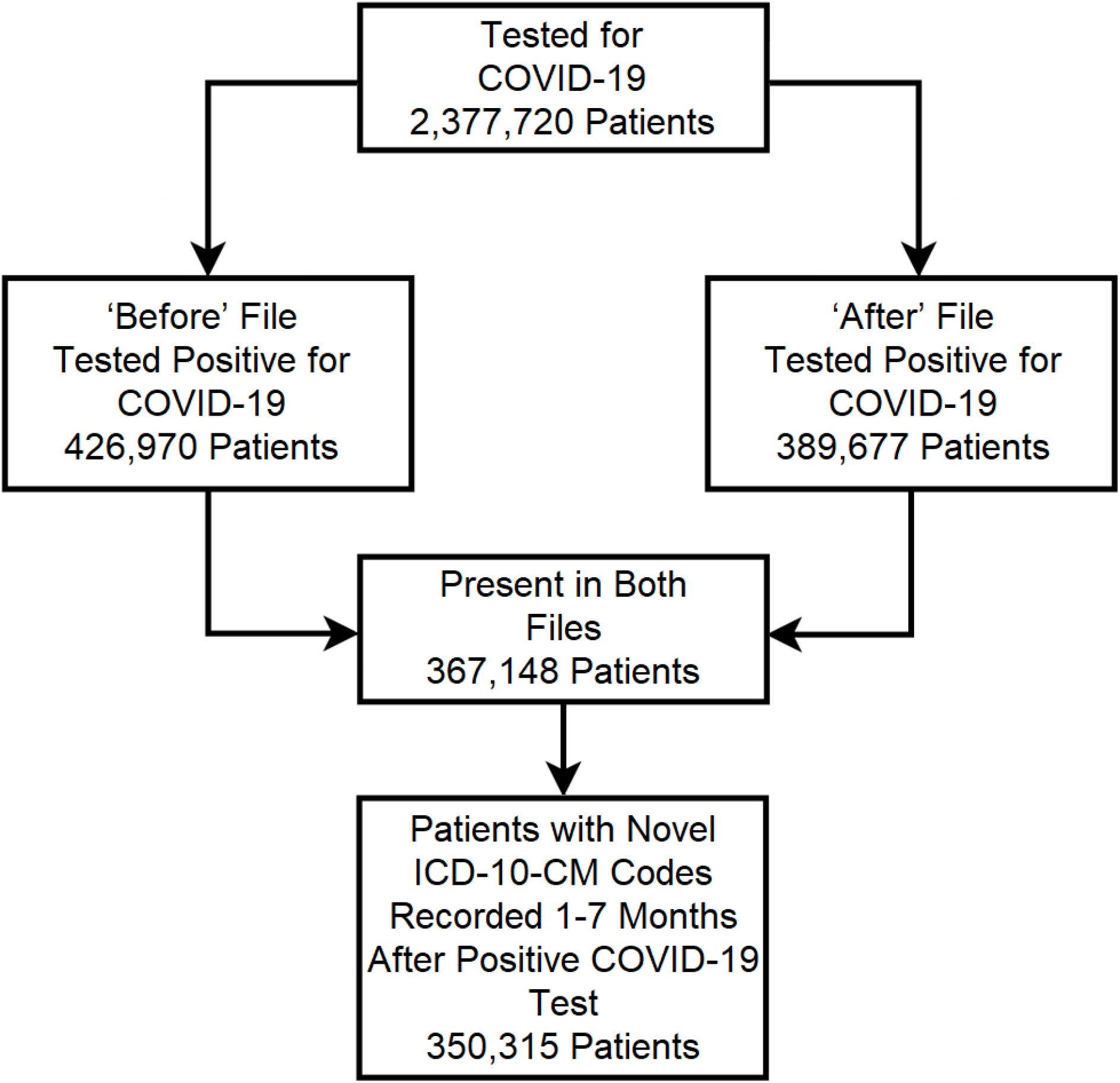
Recruitment Flowchart for the Study. We collected additional data to examine the association of demographics, comorbidities, vaccination status, and COVID-19 case severity with the incidence of long COVID. Demographic data collected included age, sex, race, and ethnicity. Comorbidities were evaluated using 2-year Elixhauser Comorbidity Indices Scores. Vaccinated patients were defined as having at least one COVID-19 vaccine dose recorded at least two weeks and no more than nine months before their positive COVID-19 test. We defined two classes of severe COVID-19 based on minimum recorded oxygen saturation. The first class, severe COVID-19, was defined by a minimum oxygen saturation of <94%.^16^ The second class, severe COVID-19 with severe desaturation, was defined by a minimum oxygen saturation of < 88%.^17^

### Data Analysis

We chose six month “before” and “after” COVID test control and case windows to allow patients to serve as their own controls. The six month “after” case window began one month after the positive COVID test. We defined “novel” ICD-10-CM codes as those that appeared in a patient’s “after” file but not in the “before” file. We calculated the frequency of each novel ICD-10-CM code as the percentage of the study cohort assigned the code. We excluded novel codes with a frequency of < 1:1000 from further analysis. We defined codes as upregulated when the frequency of that code in the after file was significantly increased as compared to the frequency in the before file, if it had a Chi-Square with a Bonferroni corrected p-value<0.00006. All resulting codes were grouped for additional analysis and organization (see ICD-10-CM Functional and Medical Specialty Groupings subsection of Methods).

We also defined ICD-10-CM functional groups as “upregulated” if they were statistically more frequent post COVID by Chi Square analysis with Bonferroni correction with a p<0.00006. We limited the long COVID ICD-10-CM code functional groups to those that were significantly increased in frequency. We used “before” and “after” frequencies to calculate odds ratios and confidence intervals for each novel ICD-10-CM functional group. We calculated odds ratios from frequency data.

We analyzed potential risk factors including vaccination status and COVID-19 severity for long COVID by creating 2×2 tables and applying Pearson Chi-Square testing. We applied a similar approach to the analysis of demographic factors. We used R version 4.1.2 and R-Studio to perform the statistical analysis.

### ICD-10-CM Functional and Medical Specialty Groupings

We grouped ICD10 CM codes in three steps. We first combined ICD-10-CM codes that had the same initial three characters. We then grouped ICD-10-CM codes with different initial characters if the diagnoses were functionally similar to create our ICD-10-CM functional groups. For example, we grouped I47.1 (supraventricular tachycardia), I47.2 (ventricular tachycardia), and R00.0 (tachycardia, unspecified) together as tachycardia. Finally, we manually curated each of these ICD-10-CM functional groups into medical specialties for organizational purposes.

### Long COVID Definition

We included in our long COVID definition each ICD-10-CM code with an incidence over six months (T_0_ + 1M – T_0_+7M)> 1:1000 and a significant overall frequency increase. Long COVID patients were defined as having any of the 323 upregulated ICD-10-CM codes between two and seven months after their positive COVID-19 diagnosis, but not in their pre-COVID diagnoses.

### Risk Factors for Long COVID

The multivariate regression models were done for each risk factor one including Age, Gender, Race, Ethnicity, 2-year Elixhauser score, a second with Age, Gender, Race, Ethnicity, 2-year Elixhauser score, O2Sat <94%, and a third with Age, Gender, Race, Ethnicity, 2-year Elixhauser score, COVID Vaccination status. We present the univariate rates as well as the results of the regression analysis using R4.1.2 and R Studio.

## Results

### Long COVID Definition

We extracted ICD-10-CM diagnosis codes assigned to 367,148 patients who underwent a positive COVID test at VA. Patients with one or more novel COVID related diagnoses numbered 268,320. The remaining 98,828 patients had no novel long COVID ICD-10-CM diagnoses in their post-COVID period when compared to their pre-COVID period. Table 1 contains the demographic characteristics of the study cohort. Males were significantly older than the females on average, 60.29 (95% CI: 95% CI: 60.24 – 60.35) versus 47.85 (95% CI: 47.73 – 47.97), respectively.

**Table 1.** Demographic data and two-year Elixhauser scores for long COVID patients and non-long COVID patients

|  | Non-Long COVID Patients (98,828) | Long COVID Patient (268,320) | P Value |
| --- | --- | --- | --- |
| Age (Mean/Conf. int.) | 52.14 (52.03 – 52.24) | 60.85 (60.79 – 60.91) | - |
| Elixhauser Score (Mean/Conf. int.) | 3.03 (2.99 – 3.07) | 7.05 (7.01 – 7.09) | - |
| Sex |  |  |  |
| Male | 75,418 (76.31%) | 234,720 (87.48%) | < 0.00001 |
| Female | 16,854 (17.05%) | 31,651 (11.80%) | < 0.00001 |
| Not Listed | 6,556 (6.63%) | 1,949 (0.73%) | < 0.00001 |
| Ethnicity |  |  |  |
| Hispanic or Latino | 10,147 (10.27%) | 26,171 (9.75%) | < 0.00001 |
| Not Hispanic or Latino | 71,595 (72.44%) | 227,404 (84.75%) | < 0.00001 |
| Not Listed | 17,086 (17.29%) | 14,745 (5.50%) | < 0.00001 |
| Race |  |  |  |
| American Indian or Alaska Native | 743 (0.75%) | 2,243 (0.84%) | 0.01182 |
| Asian | 1,420 (1.44%) | 2,973 (1.11%) | < 0.00001 |
| Black or African American | 20,779 (21.03%) | 65,218 (24.31%) | < 0.00001 |
| Native Hawaiian/Other Pacific Islander | 905 (0.92%) | 2,522 (0.94%) | 0.49900 |
| White | 55,359 (56.02%) | 173,169 (64.54%) | < 0.00001 |
| Not Listed | 19,622 (19.85%) | 22,195 (8.27%) | < 0.00001 |

We developed a definition of long COVID consisting of 323 ICD-10-CM diagnosis codes grouped into 143 ICD-10-CM functional groups that were significantly increased in our 367,148 patient post-COVID population. We define seventeen medical specialty long COVID subtypes including cardiology long COVID, neurology long COVID, and pulmonary long COVID. Table 2 shows the ICD-10-CM functional groups and medical specialties. Within each field, the ICD-10-CM code groups are sorted in descending order by their Odds Ratios.

**Table 2.** Long COVID definition results by ICD-10-CM Functional group description and medical specialty.

| Functional Group Description | Medical Specialty | ICD-10-CM Codes | Novel Count | Odds Ratio |
| --- | --- | --- | --- | --- |
| Hypotension | Cardiology | I95.1, I95.89, I95.9 | 7865 | 1.271538 |
| Heart Failure Diastolic Dysfunction | Cardiology | I11.0 | 4594 | 1.26876 |
| Cardiac Arrhythmia | Cardiology | I47.1, I48.0, I48.19, I48.20, I48.21, I48.3, I48.91, I48.92, I49.3, R00.0, R00.2 | 35648 | 1.223517 |
| Cardiovascular Disease | Cardiology | I21.A1, I24.8, I25.10, I25.2, I25.5, I42.8, I42.9, I50.20, I50.22, I50.23, I50.30, I50.32, I50.33, I50.42, I50.9, I71.40 | 45157 | 1.206416 |
| Valve Stenosis/Insufficiency | Cardiology | I27.82, I34.0, I35.0 | 3916 | 1.171221 |
| Peripheral Vascular Disease | Cardiology | I73.9 | 4862 | 1.105507 |
| Hyperlipidemia | Cardiology | E78.2, E78.5 | 54913 | 1.070793 |
| Hypertension | Cardiology | I10 | 39677 | 1.046517 |
| Gingival Recession | Dentistry | K06.010, K06.020 | 2497 | 1.256852 |
| Gingivitis | Dentistry | K03.0, K05.00, K05.10, K05.321, K05.322, K06.1 | 7818 | 1.235098 |
| Dental Caries | Dentistry | K02.3, K02.51, K02.52, K02.62, K02.7 | 19887 | 1.154138 |
| Pressure ulcer | Dermatology | L89.152, L89.153, L89.159 | 1639 | 2.3395 |
| Nonscarring Hair Loss | Dermatology | L65.9 | 441 | 1.853409 |
| Melanocytic Nevi | Dermatology | D22.9 | 1886 | 1.195428 |
| Nail Dystrophy | Dermatology | L60.3 | 5209 | 1.152928 |
| Xerosis Cutis | Dermatology | L85.3 | 4048 | 1.133044 |
| Fatigue | Endocrinology | G93.3 | 542 | 10.18039 |
| Malnutrition | Endocrinology | E43, E44.0, E46, E63.9, R63.8, R64 | 6372 | 1.856287 |
| Adult Failure to Thrive | Endocrinology | R62.7 | 1827 | 1.578271 |
| Phosphorus Metabolism Disorder | Endocrinology | E83.39 | 1306 | 1.297077 |
| Abnormal Weight Loss | Endocrinology | R63.4 | 4439 | 1.16755 |
| Osteoporosis | Endocrinology | M81.0 | 1442 | 1.120668 |
| Prediabetes | Endocrinology | R73.03 | 6402 | 1.105807 |
| Type 2 Diabetes Mellitus | Endocrinology | E11.22, E11.3291, E11.36, E11.40, E11.42, E11.51, E11.649, E11.65, E11.9 | 49454 | 1.104288 |
| Hypothyroidism | Endocrinology | E03.9 | 7059 | 1.050629 |
| Obesity | Endocrinology | E66.9 | 19041 | 1.038481 |
| Ileus | Gastroenterology | K56.7 | 693 | 1.585583 |
| Enterocolitis | Gastroenterology | A04.72 | 760 | 1.568205 |
| Elevation of Liver Transaminase Levels | Gastroenterology | R74.01 | 2117 | 1.515881 |
| Polyp of Stomach and Duodenum | Gastroenterology | K31.7 | 718 | 1.339522 |
| Ascites | Gastroenterology | R18.8 | 795 | 1.332139 |
| Esophagitis | Gastroenterology | K08.499, K20.80, K20.90, K22.89 | 3013 | 1.317627 |
| Portal Hypertension | Gastroenterology | K76.6 | 671 | 1.291358 |
| Dysphagia | Gastroenterology | R13.10, R13.11, R13.12, R13.13 | 13011 | 1.274248 |
| Constipation | Gastroenterology | K59.00, K59.03, K59.09 | 13376 | 1.224596 |
| Diseases of Stomach and Duodenum | Gastroenterology | K31.89 | 1394 | 1.193473 |
| Gastritis | Gastroenterology | K29.50, K29.70 | 2963 | 1.191019 |
| Gastrointestinal Hemorrhage | Gastroenterology | K92.2 | 2320 | 1.186902 |
| Diverticulosis | Gastroenterology | K57.30 | 5522 | 1.168767 |
| Melena | Gastroenterology | K92.1 | 2334 | 1.143609 |
| Cirrhosis of Liver | Gastroenterology | K74.60 | 1683 | 1.13734 |
| Gastro-Esophageal Reflux Disease | Gastroenterology | K21.00, K21.9 | 31020 | 1.086091 |
| Fatty Liver | Gastroenterology | K76.0 | 4912 | 1.081379 |
| Homelessness | General Internal Medicine | Z59.00, Z59.01, Z59.02, Z59.811, Z59.812, Z59.819, Z59.89 | 11511 | 1.456793 |
| Vitamin Deficiency | General Internal Medicine | E53.8, E55.9 | 21619 | 1.105953 |
| Pulmonary Embolism without Acute Cor Pulmonale | Hematology | I26.99 | 2670 | 1.804971 |
| MGUS | Hematology | E88.09, R77.8 | 1273 | 1.542297 |
| Acute Embolism and Thrombosis | Hematology | I82.401, I82.402, I82.409, I82.890 | 2510 | 1.42562 |
| Pancytopenia | Hematology | D61.818 | 939 | 1.291209 |
| Thrombocytopenia | Hematology | D69.59, D69.6 | 3876 | 1.260737 |
| Abnormal Coagulation Profile | Hematology | R79.1 | 826 | 1.250246 |
| Anemia | Hematology | D50.0, D50.9, D53.9, D62, D63.0, D63.1, D63.8, D64.89, D64.9 | 30750 | 1.242379 |
| Abnormal White Blood Cell Count | Hematology | D72.829 | 3528 | 1.167524 |
| Infectious Sequelae | Infectious Diseases | B94.8 | 606 | 23.42175 |
| Viral Pneumonia | Infectious Diseases | J12.89 | 1604 | 2.478623 |
| Influenza | Infectious Diseases | J09.X2 | 547 | 2.193782 |
| Thrush | Infectious Diseases | B37.0 | 537 | 1.844434 |
| Sepsis | Infectious Diseases | A41.89, A41.9, R65.20, R65.21 | 6816 | 1.608247 |
| Pseudomonas | Infectious Diseases | B96.5 | 682 | 1.595698 |
| Enterococcus | Infectious Diseases | B95.2 | 884 | 1.594505 |
| Bacterial Pneumonia | Infectious Diseases | J15.9 | 2862 | 1.54475 |
| MRSA | Infectious Diseases | B95.62, Z22.322 | 1316 | 1.488166 |
| Bacteremia | Infectious Diseases | R78.81 | 2139 | 1.377602 |
| Klebsiella | Infectious Diseases | B96.1 | 728 | 1.376463 |
| MSSA | Infectious Diseases | B95.61, B95.7 | 1015 | 1.341265 |
| Proteus | Infectious Diseases | B96.4 | 511 | 1.336283 |
| E. Coli | Infectious Diseases | B96.20 | 1077 | 1.319389 |
| Osteomyelitis | Infectious Diseases | M86.9 | 1121 | 1.224393 |
| Urinary Tract Infection | Infectious Diseases | N39.0, T83.511A | 8553 | 1.161612 |
| Drug Toxicity | Nephrology | T38.0X5A, T45.1X5A | 1073 | 1.686641 |
| Dehydration | Nephrology | E86.1 | 959 | 1.293327 |
| Acute Kidney Failure | Nephrology | N17.0, N17.8, N17.9 | 11392 | 1.290399 |
| Disorders of Fluid, Electrolyte and Acid-Base Balance | Nephrology | E83.42, E83.52, E87.0, E87.1, E87.2, E87.20, E87.3, E87.4, E87.5, E87.6, E87.70, E87.8 | 25885 | 1.276923 |
| Chronic Kidney Disease | Nephrology | I12.0, I12.9, I13.0, I13.2, N18.2, N18.30, N18.31, N18.32, N18.4, N18.6, N18.9, N28.89, Z99.2 | 35590 | 1.258035 |
| Edema | Nephrology | R60.0, R60.9 | 11683 | 1.116561 |
| Encephalopathy | Neurology | G92.8, G93.40, G93.41, G93.49 | 4476 | 1.705173 |
| Delirium | Neurology | F05 | 1414 | 1.639297 |
| Lower Back Pain | Neurology | M54.50 | 24694 | 1.578581 |
| Muscle Weakness | Neurology | M62.81 | 9362 | 1.277051 |
| Need for Assistance with Personal Care | Neurology | Z74.1 | 3287 | 1.276263 |
| Weakness | Neurology | R53.1, R54 | 15912 | 1.258534 |
| Cognitive Impairment | Neurology | R41.0, R41.81, R41.82, R41.841, R41.89, R41.9 | 10045 | 1.241561 |
| Wheelchair | Neurology | Z99.3 | 654 | 1.234073 |
| Seizure Disorder | Neurology | G40.909 | 1155 | 1.205124 |
| Dementia | Neurology | F01.50, F02.80, F03.90, G30.9, G31.84 | 10406 | 1.204819 |
| Falls | Neurology | R29.6 | 1879 | 1.199129 |
| Difficulty in Walking | Neurology | R26.2, R26.89, R26.9, Z74.09 | 22449 | 1.171356 |
| Falling | Neurology | Z91.81 | 3164 | 1.157633 |
| Tinnitus | Neurology | H93.13 | 8889 | 1.135359 |
| Cerebrovascular Disease | Neurology | I63.9, I69.351, I69.354 | 4200 | 1.119552 |
| Neuropathy | Neurology | G62.9, G89.18, G89.29, G89.3, G89.4 | 23681 | 1.118604 |
| Cervicalgia | Neurology | M54.2 | 13691 | 1.052616 |
| Malignant Neoplasm of Liver | Oncology | C78.7 | 420 | 1.704389 |
| Malignant Neoplasm of Lung | Oncology | C34.90 | 657 | 1.350535 |
| Malignant Neoplasm of Bone | Oncology | C79.51 | 622 | 1.314306 |
| Solitary Pulmonary Nodule | Oncology | R91.1 | 5557 | 1.116382 |
| Conjunctivitis | Ophthalmology | H10.45 | 2645 | 1.180325 |
| Retinopathy | Ophthalmology | H35.372, H35.373 | 2413 | 1.173309 |
| Dry Eye Syndrome | Ophthalmology | H04.123 | 19581 | 1.164934 |
| Presbyopia | Ophthalmology | H52.4 | 36752 | 1.154351 |
| Myopia | Ophthalmology | H52.13 | 8012 | 1.151422 |
| Vitreous Degeneration | Ophthalmology | H43.813 | 3085 | 1.142235 |
| Astigmatism | Ophthalmology | H52.203, H52.223 | 13737 | 1.140477 |
| Unspecified Disorder of Refraction | Ophthalmology | H52.7 | 7093 | 1.126198 |
| Open Angle with Borderline Findings | Ophthalmology | H40.013 | 6278 | 1.121644 |
| Cataracts | Ophthalmology | H25.012, H25.11, H25.12, H25.13, H25.811, H25.812, H25.813, H26.9 | 42904 | 1.119152 |
| Hypermetropia | Ophthalmology | H52.03 | 7317 | 1.110326 |
| Epistaxis | Otolaryngology | R04.0 | 1342 | 1.235389 |
| Dysphonia | Otolaryngology | R49.0 | 1368 | 1.182724 |
| Impacted Cerumen | Otolaryngology | H61.23 | 3780 | 1.121918 |
| Sensorineural Hearing Loss | Otolaryngology | H90.3, H90.A22 | 17093 | 1.1138 |
| Restlessness and Agitation | Psychiatry / Psychology | R45.1 | 893 | 1.279866 |
| Mental Disorder | Psychiatry / Psychology | F09, F32.A, F41.1, F41.9, F43.12 | 48256 | 1.135075 |
| Malaise | Psychiatry / Psychology | R53.81 | 2251 | 1.132321 |
| Problems Related to Psychosocial Circumstances | Psychiatry / Psychology | Z65.8, Z65.9 | 23892 | 1.102123 |
| Acute Respiratory Distress Syndrome | Pulmonary | J80 | 688 | 5.299347 |
| Dependence on Ventilator | Pulmonary | Z99.11 | 427 | 3.146519 |
| Chronic Cough | Pulmonary | R05.3 | 2113 | 2.026076 |
| Respiratory Failure | Pulmonary | J96.00, J96.01, J96.02, J96.10, J96.11, J96.20, J96.21, J96.22, J96.90, J96.91 | 14882 | 1.997749 |
| Dependence on Supplemental Oxygen | Pulmonary | R09.02, Z99.81 | 7792 | 1.715279 |
| Interstitial Pulmonary Disease | Pulmonary | J84.9 | 685 | 1.536233 |
| Pulmonary Fibrosis | Pulmonary | J84.10 | 1232 | 1.527598 |
| Pleural Effusion | Pulmonary | J90, J91.8 | 2990 | 1.4413 |
| Abnormalities of Breathing | Pulmonary | R06.89 | 1107 | 1.338466 |
| Pneumonia | Pulmonary | J16.8, J17, J18.8, J18.9, J69.0 | 8181 | 1.315208 |
| Acute Pulmonary Edema | Pulmonary | J81.0 | 624 | 1.295683 |
| Pulmonary Hypertension | Pulmonary | I27.20 | 1782 | 1.282603 |
| Respiratory Disorders | Pulmonary | J98.4, J98.8, J98.9 | 3711 | 1.281901 |
| Emphysema | Pulmonary | J43.9 | 1700 | 1.229811 |
| Dyspnea | Pulmonary | R06.00, R06.02, R06.03, R06.09 | 27926 | 1.206525 |
| Atelectasis | Pulmonary | J98.11 | 1766 | 1.206262 |
| Diaphragmatic Hernia | Pulmonary | K44.9 | 2952 | 1.169074 |
| Chronic Obstructive Pulmonary Disease | Pulmonary | J44.0, J44.9 | 12639 | 1.131736 |
| Asthma | Pulmonary | J45.909 | 4126 | 1.093696 |
| Sleep Disorders | Pulmonary | G47.00, G47.33 | 41147 | 1.090759 |
| Osteoarthritis | Rheumatology | M17.0 | 4546 | 1.090886 |
| Gout | Rheumatology | M10.9 | 5997 | 1.057874 |
| Uropathy | Urology | N13.9 | 597 | 1.256139 |
| Retention of Urine | Urology | R33.8, R33.9 | 6269 | 1.251816 |
| Obstructive and Reflux Uropathy | Urology | N13.8 | 1385 | 1.204741 |
| Urinary Incontinence | Urology | N31.9, R32 | 3843 | 1.153016 |
| Benign Prostatic Hyperplasia | Urology | N40.0, N40.1 | 24210 | 1.128437 |
| Frequency of Micturition | Urology | R35.0 | 3032 | 1.106673 |

Figures 3 and 4 show the signs, symptoms, and diagnoses with significantly increased relative risks in the post-COVID period with their respective confidence intervals sorted by medical specialty.

**Figure 3.**
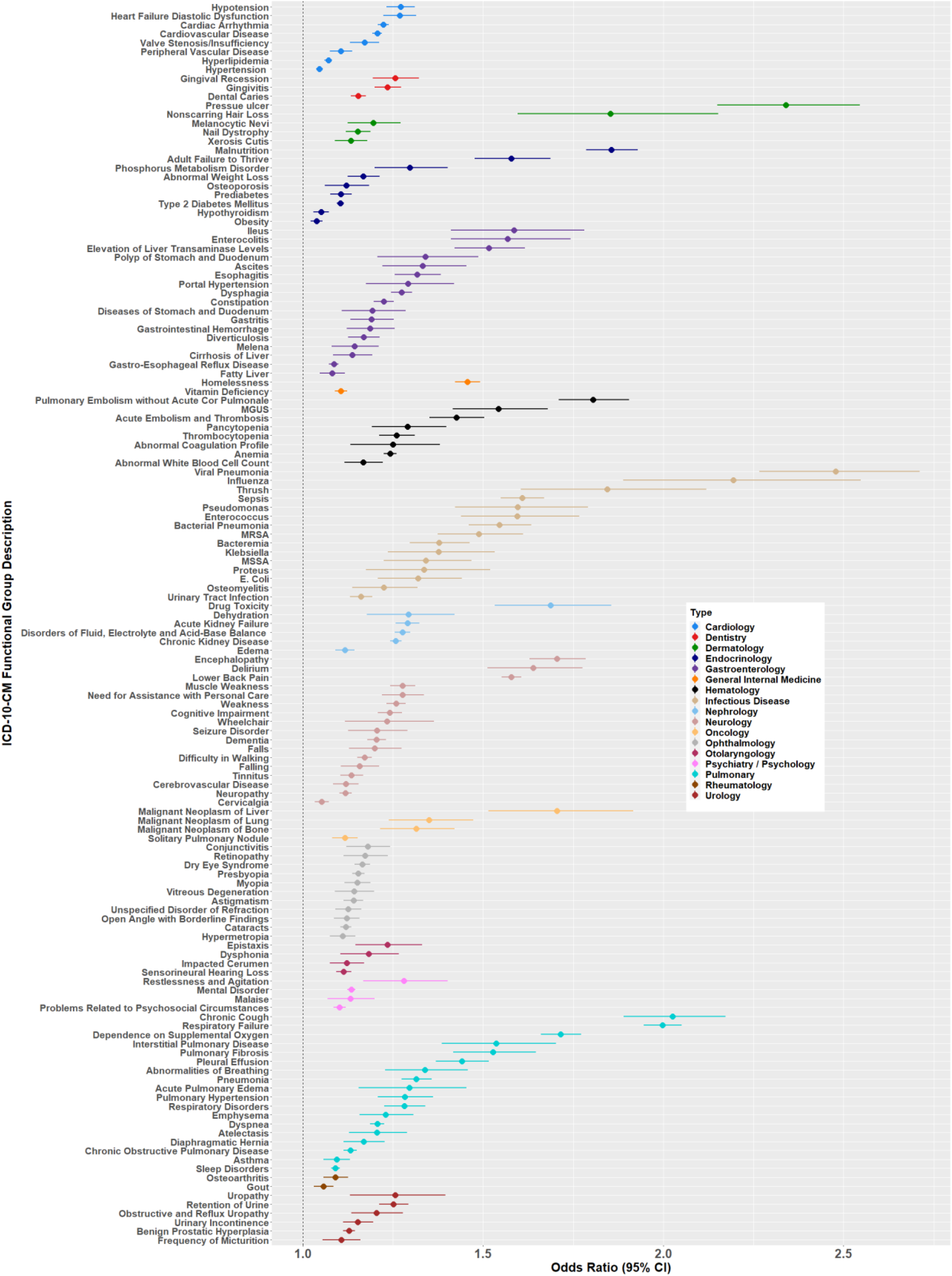
Odds ratios < 3 for long COVID ICD-10-CM functional groups by medical specialty subtype.

**Figure 4.**
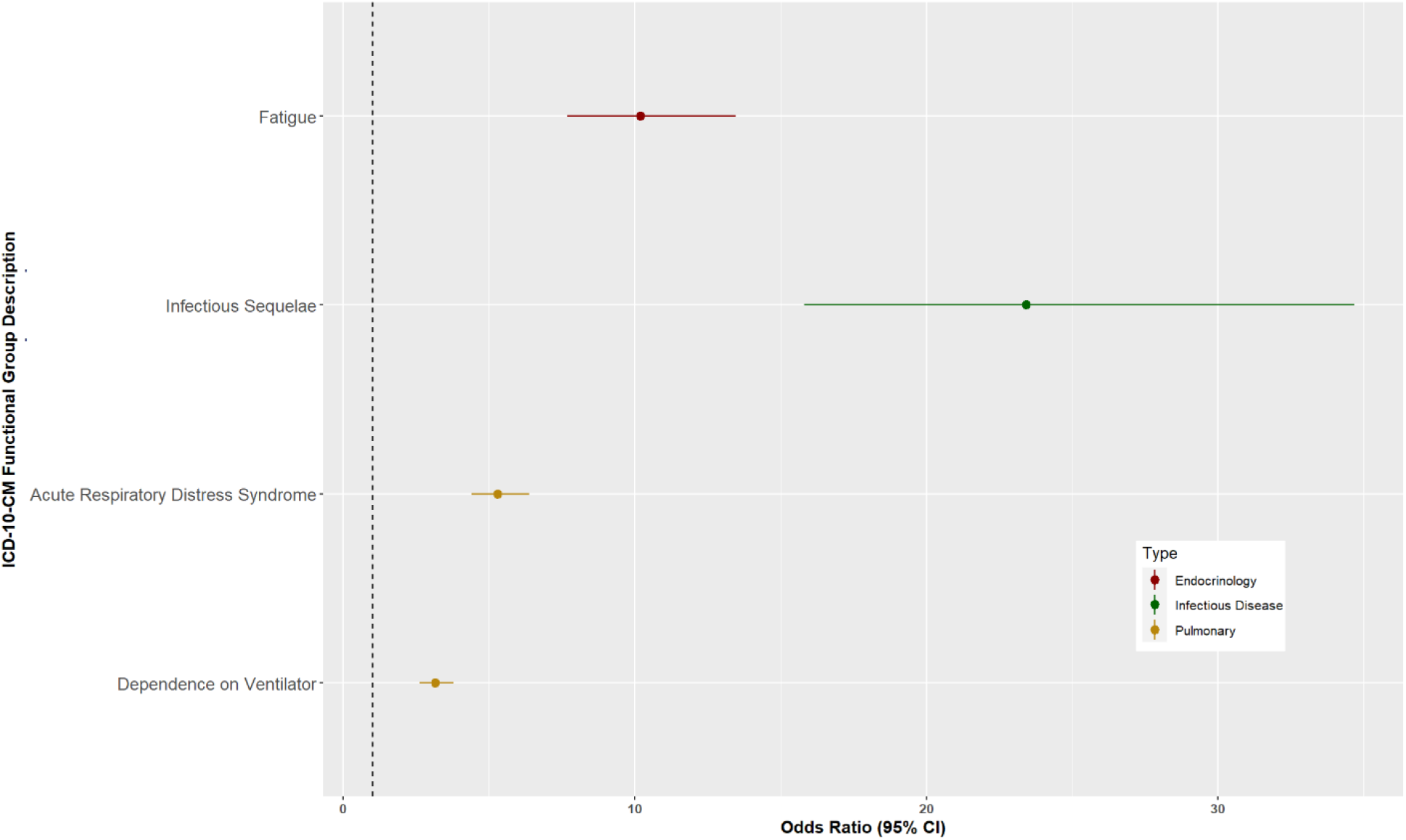
Odds ratios > 3 for long COVID ICD-10-CM functional groups by medical specialty subtype.

Case counts were greatest for the specialties of Cardiology (196,6320, Neurology (159,358), Ophthalmology (149,817) and Pulmonary (138,470). The lowest case counts were for Oncology (7,256), Rheumatology (10,543) and Dermatology (13,233).

COVID-19 test positive patients were assigned novel signs, symptoms, or diagnoses included in our definition of long COVID at a rate of between 59.7% (percentage based on COVID positive patients tested at the VA) and 76.6% (percentage based on all COVID positive patients with diagnostic history and follow up diagnoses one to seven months after test).

Most long COVID patients were documented with at least one ICD-10-CM code found in our long COVID definition within three months of their positive COVID-19 test. The percentage of patients documented with their first long COVID ICD-10-CM code decreases with each subsequent month (See Figure 6).

**Figure 5.**
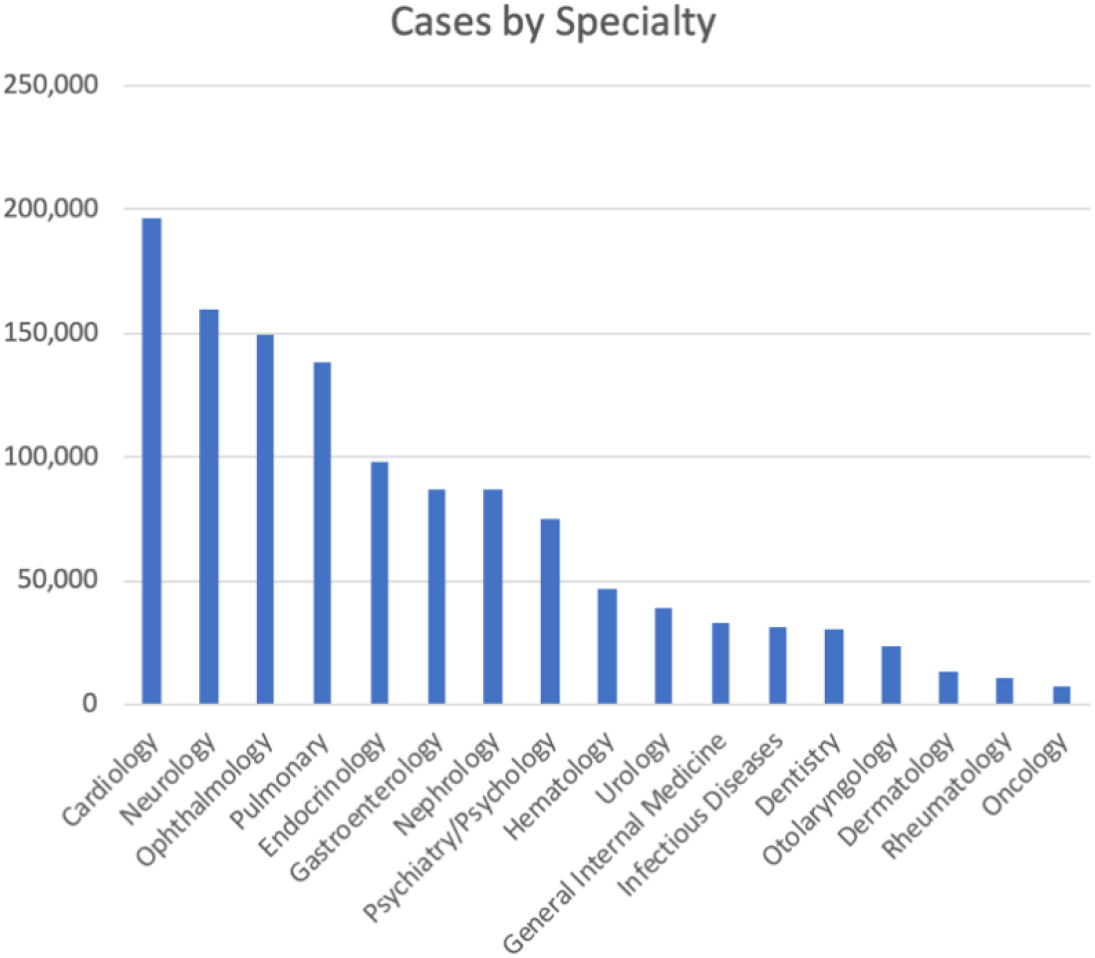
Case counts by medical specialty.

**Figure 6.**
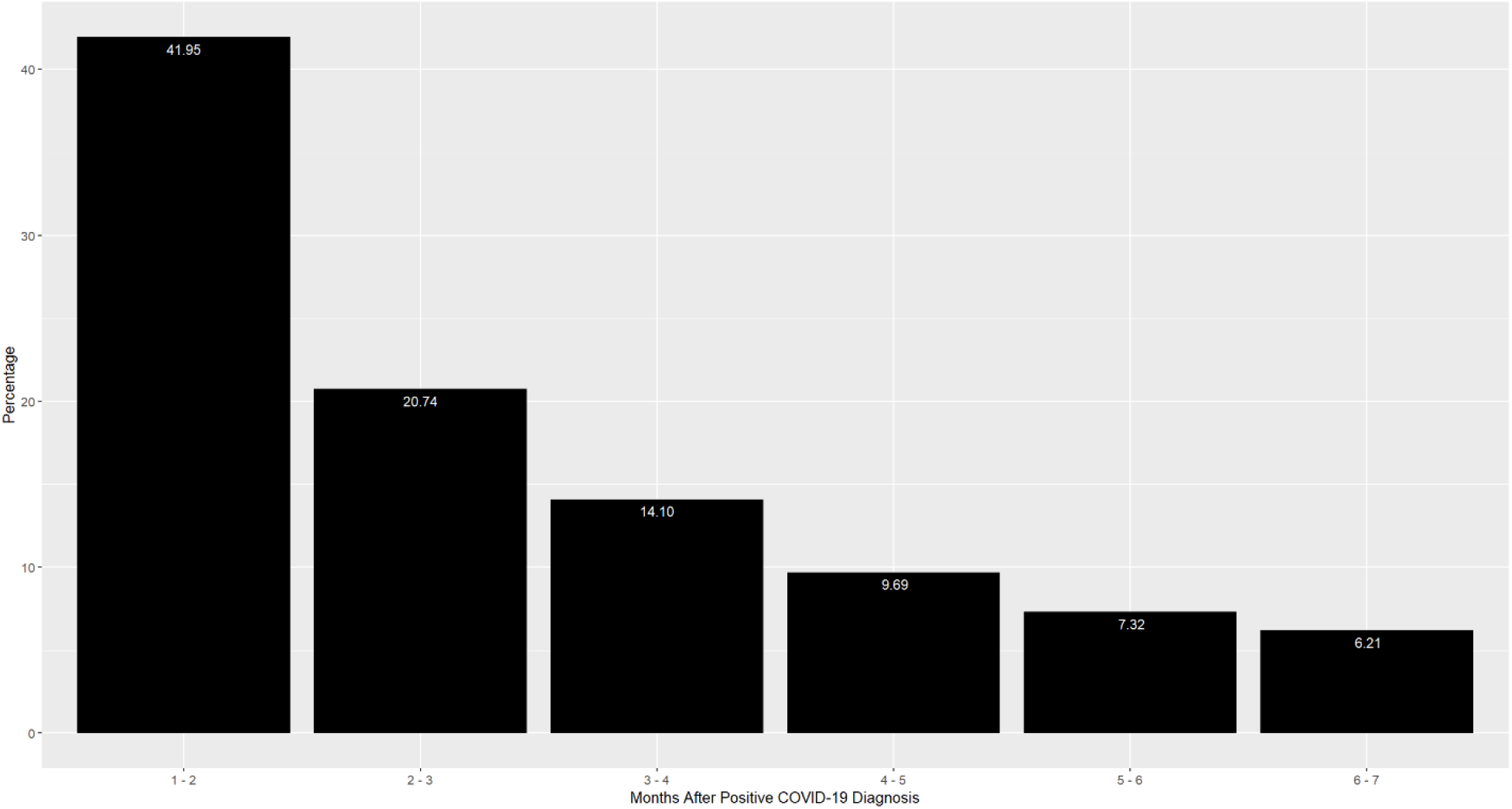
Percentage of prospective post COVID-19 diagnosis (Months 1-7) that each long COVID patient had their first long COVID ICD-10-CM code documented. Month 0-1 ICD-10-CM codes were not included as they included acute symptoms.

### Risk Factors for Long COVID

We presented in Table 1 a comparison of demographic characteristics and Elixhauser comorbidity scores of long COVID patients and non-long COVID patients. The long COVID cohort was older with more comorbidities. The long COVID cohort also had higher percentages of White and Black individuals and non-Hispanic and non-Latino ethnicities. Patients with a 2-year Elixhauser score of greater than 21 had a much higher proportion developing long COVID (p < 0.001, Pearson Chi-Square) (See Table 3).

**Table 3.** Proportion of patients that developed long COVID comparing different 2-year Elixhauser score ranges.

| 2 Year Elixhauser Score | No Long COVID Count | Long COVID Count | Percent Long COVID (95% CI) |
| --- | --- | --- | --- |
| 0-21 | 96,055 | 242,108 | 71.60% (71.44% - 71.75%) |
| 22-42 | 2,454 | 22,716 | 90.25% (89.88% - 90.61%) |
| 43-63 | 308 | 3,317 | 91.50% (90.55% - 92.36%) |
| 64-84 | 11 | 179 | 94.21% (89.93% - 96.77%) |

Our data did not indicate that vaccination was protective against the development of long COVID. However, vaccination resulted in significantly lower rates of novel ARDS in the post-covid period (13.2% CI: 10.4%-16.9%) as compared with the unvaccinated population (19.6% CI: 18.1% - 21.2), p<0.001.

Patients with minimum O2 saturations constituting severe COVID and severe COVID with severe desaturation were significantly more likely to develop long COVID (both had p-values<0.001, Pearson Chi-Square) (See Table 4).

**Table 4.** Low oxygen saturations and the proportion of patients that developed long COVID.

| Severe COVID | No Long COVID Count | Long COVID Count | Percent Long COVID (95% CI) |
| --- | --- | --- | --- |
| Low O2 (NIH Definition*) | 3,903 | 29,411 | 88.28 % (87.93% - 88.63%) |
| No Low O2 (NIH Definition*) | 94,925 | 238,909 | 71.57% (71.41% - 71.72%) |
| Low O2 (Severe Desaturation**) | 637 | 5,566 | 89.73% (88.95% - 90.46%) |
| No Low O2 (Severe Desaturation**) | 98,191 | 262,754 | 72.80% (72.65% - 72.94%) |
| * Min(O2 Sat) < 94% ** Min(O2 Sat) < 88% |  |  |  |

The multivariate regression models all confirmed that COVID patients during the Omicron variant predominant period were at slightly higher risk of developing Long COVID at p-value < 0.001.

## Discussion

Numerous reports document specialty-specific signs, symptoms and diagnoses correlated with long COVID. We present a novel analysis based on a large national data set and the full multispecialty breadth of ICD-10-CM diagnosis codes to create an overall holistic long COVID definition that confirms and extends previous reports.

We allowed patients to be their own controls and used the entire cohort before and after COVID-19 infection to determine the relative risk of signs, symptoms, and disorders. This ensured that the signal was both novel and upregulated. We found COVID-19 positive patients developed signs, symptoms, or diagnoses included in our long COVID definition at a proportion of between 59.7% (percentage based on COVID positive patients tested at the VA) and 76.6% (percentage based on all COVID positive patients with diagnostic history and follow up diagnoses one to seven months after test). More than three-fourths of long COVID patients met our long COVID definition within four months of their positive COVID-19 test.

We found long COVID frequency differences based on race and ethnicity. These differences may be related to socioeconomic status, which is directly correlated with the presence of comorbidities.^18–20^ The long COVID cohort was eight years older with more comorbidities (two-year Elixhauser score 7.97 in the long Covid patients vs 4.21 in the non-long Covid patients). In our cohort, the males were significantly older than the females on average, 60.29 (95% CI: 95% CI: 60.24 – 60.35) versus 47.85 (95% CI: 47.73 – 47.97), respectively. We found that long COVID frequency was increased in female patients, the more severely ill, and patients who had a more severe bout of COVID as judged by their minimum oxygen saturation.

We found 143 upregulated diagnostic groups, with odds ratios as high as 23. We also found seventeen upregulated medical specialty groupings containing between three and twenty-one signs, symptoms, or diagnoses. This provides strong evidence for a broad definition of long COVID.

Carfi et al. found that most common long-term symptoms were fatigue, dyspnea, joint pain, and chest pain.^2^ Each except joint pain is represented in our long COVID definition. However, joint pain may be related to findings in our definition such as difficulty walking and an overall decrease in mobility. COVID-19 is known to cause lung abnormalities, especially in cases with pneumonia.^21^ We found that the likelihood of developing pneumonia after COVID-19 infection is significantly upregulated, potentially interconnected with the numerous findings in our Pulmonary long COVID definition. Autopsy evaluation of COVID-19 victims’ lung tissue demonstrated diffuse alveolar damage with perivascular T-cell infiltration and severe endothelial injury. ^22^ Long COVID patients have been found to have abnormal ^129^Xe MRI gas exchange and CT vascular density measurements, which we postulate could be related to the pulmonary fibrosis (J84.10) or emphysema (J43.9) diagnoses identified in our definition. ^23^

Our definition shows that the long-term effects of COVID-19 are associated with damage to numerous body systems including the kidneys, heart, eyes, and nervous system. Our results are corroborated by other studies. Cognitive dysfunction (brain fog) is often associated with long COVID and can be difficult to diagnose and treat.^5^ COVID-19 infection is far more likely to cause cardiac complications than vaccination.^24^ The gastrointestinal codes we observed reflect previous literature^25^ and may relate to reported alterations to the gastrointestinal tract after COVID-19.^3,4^ Finally, previous studies have noted that COVID can alter ocular physiology, supporting our ophthalmology related findings.^26^

Patients with more severe cases of COVID 19, as manifest by low oxygen saturations, should be watched carefully for the development of long COVID as they were significantly more likely to develop long COVID. Sicker patients with higher 2-year Elixhauser scores were significantly more likely to develop long COVID. Patients with multiple comorbidities should be made aware of this risk and participate in active surveillance for the development of signs and symptoms of long COVID.

The American Medical Association (AMA) notes there are three categories of long COVID patients: Those who do not recover completely and have ongoing symptoms; those with symptoms related to chronic hospitalization; and those who develop new symptoms after recovery.^27^ In our study, we did not differentiate by these subtypes and instead leave that to future research. It is possible that some of these signs and symptoms may have occurred during the first month and may be the persistent subtype. It is possible that some of the upregulated codes may be found with other serious illnesses, though only 9.1% of our cohort had severe COVID based on oxygen saturation <94%. We are not able to distinguish conditions that represent acceleration of pre-existing disease from those that represent de novo COVID-related conditions. For example, is the increased incidence of Non-ST elevation (NSTEMI) myocardial infarction (I21.4) related to the general stress of acute illness impacting preexisting coronary artery disease or to an underlying de novo long COVID related condition? Better understanding will require additional research. In any event, whether causal or associative, de novo disease or exacerbation of chronic disease, new or persistent clinical problems require assessment, treatment, and monitoring.

Limitations include that the cohort study population is 84% male, reflective of the overall VA patient population which is between 87% and 95% male (depending upon data source and whether gender has been self-reported).^28,29^ Additionally, the male Veteran population who use the VA healthcare system is older than the population of female Veterans who use VA. Our study did not include home testing for Covid-19 that went unreported to the VA healthcare system. Patients who tested positive during the omicron dominant time period were slightly more likely to develop Long COVID when compared to the earlier strains (76% vs 72%, p<0.001). The reality of emerging viral variants emphasizes the need for a well-defined and well-maintained definition of long COVID over time and with variant-specific derivation. The study was not powered to show independence of the individual risk factors for long COVID.

In this case-crossover study, we hope that our empirically defined long COVID definition will lead to more consistent identification of long COVID and its medical specialty subtypes and support of a variety of COVID related initiatives. Our definition is actionable as individuals who have multiple co-morbidities and more severe bouts of COVID should be followed more closely for the development of long covid signs or symptoms. Our definition can also inform screening questions for high-risk patients. For example, helping clinicians identify patients with enhanced long COVID risk who may benefit from monitoring programs or patients with previously undiagnosed long COVID for whom it may be appropriate to create a referral to a long COVID clinic. We also anticipate that our long COVID definition may support through standardization of future subspecialty specific long COVID research.

Future research should look at health outcomes for each long COVID medical specialty subtypes to identify those at greatest risk of developing severe morbidity. Predictive analytics should be employed to help refer these individuals earlier to monitoring and treatment programs.

As of March 5^th^, 2023, there have been 759 million confirmed cases of COVID-19 worldwide.^30^ Case counts are ever increasing. As Dr. Levine notes, immediately useful long COVID definitions are needed as are ultimately more fully inclusive definitions. ^14^ We offer our long COVID definition as a public health contribution to our pandemic response.

## Data Availability

The data can be made available to VA researchers with IRB approval.

## Acknowledgements

This work has been supported in part by grants from NIH NLM T15LM012495, R25LM014213, NIAAA R21AA026954, R33AA0226954 and NCATS UL1TR001412. This study was funded in part by the Department of Veterans Affairs.

